# Ultrasound-Guided Canggui Tanxue Acupuncture for Post-Paralytic Facial Synkinesis: A Randomized Controlled Trial

**DOI:** 10.1101/2025.07.02.25330710

**Authors:** Yuan Zhou, Xiaoming Wu, Xi Zhou, Linjia Wang, Jingxin Mao, Yanru Wang

## Abstract

**Objective:** This randomized controlled trial aimed to evaluate the efficacy and safety of ultrasound-guided Canggui Tanxue acupuncture in treating post-paralytic facial synkinesis.

**Methods:** Sixty-four patients were randomly allocated to either an Ultrasound-guided Group (n=31) receiving real-time shear wave elastography (SWE)-navigated Canggui Tanxue acupuncture targeting hypertonic muscles (Levator Labii Superioris, Depressor Anguli Oris, Depressor Labii Inferioris) with Young’s modulus >10 kPa, or a Control Group (n=33) receiving conventional meridian-based acupuncture with facial "three-line" method. Both groups received six treatment sessions over three weeks. Primary outcome was SWE-quantified reduction in Young’s modulus of target muscles. Secondary outcomes included House-Brackmann (H-B) grading, Facial Disability Index (FDI), and clinical efficacy rate based on H-B improvement.

**Results:** The Ultrasound-guided Group demonstrated significantly greater reductions in Young’s modulus versus controls (Depressor Anguli Oris: Δ3.00 kPa vs. 1.10 kPa, P=0.001; Levator Labii Superioris: Δ5.60 kPa vs. 1.00 kPa, P<0.001). Clinically, the intervention group achieved superior facial functional recovery with an 87.1% total efficacy rate compared to 42.4% in controls (P<0.001), alongside significantly better improvement in FDI physical function scores (74.84±8.42 vs. 68.94±13.68, P=0.04). However, no significant intergroup difference was observed in FDI social function scores (FDIS: P=0.92), suggesting psychosocial recovery may lag behind functional improvements. Only two minor subcutaneous hematomas (6.5%) were observed with no neurovascular complications.

**Conclusion:** Ultrasound-guided Canggui Tanxue acupuncture significantly reduces pathological muscle tension and improves functional outcomes in post-paralytic synkinesis, demonstrating its potential as a safe and precise treatment modality.

## 1 INTRODUCTION

Post-paralytic facial synkinesis is a debilitating sequela of facial paralysis characterized by involuntary, abnormal co-contraction of facial muscles during voluntary movements, often manifesting as paradoxical deviation of the oral commissure towards the affected side during the later stages of facial paralysis[1, 2]. This condition typically arises from prolonged unresolved facial muscle paralysis, where muscles transition from a flaccid state to a stiff and spastic condition. It can lead to functional impairments in eating, drinking, and facial expression control, along with disfiguring appearance, significantly compromising patients’ psychological well-being and quality of life[3, 4]. Its pathological mechanisms remain incompletely elucidated but may involve aberrant reinnervation, where regenerating nerve fibers erroneously enter adjacent pathways to misinnervate effectors, or fibrosis of affected facial muscles and abnormal increases in muscle tension following neural regeneration[5].

Current mainstay treatments for facial synkinesis, such as botulinum toxin injections, offer symptomatic relief but are transient, require repeated administrations, and do not address the underlying biomechanical abnormalities. Pharmacological interventions (e.g., corticosteroids), physical therapies (e.g., laser, infrared radiation), and surgical approaches (e.g., facial nerve decompression, botulinum toxin injections) may partially alleviate neural inflammation and edema[6-10]. However, they are generally associated with transient effectiveness, significant side effects, or surgical risks, resulting in suboptimal patient compliance[11, 12]. While microvascular decompression represents a potentially curative option, it frequently entails postoperative inefficacy, recurrence, and complications such as hearing impairment[13]. Consequently, exploring safer and more effective therapeutic strategies is imperative.

Acupuncture demonstrates potential in managing sequelae of facial paralysis. Substantial clinical evidence confirms that acupuncture can facilitate facial nerve functional recovery and ameliorate symptoms such as facial muscle spasms and synkinesis[4]. Diverse techniques, including conventional acupuncture (e.g., Chifeng Yingyuan Acupuncture), electroacupuncture, fire acupuncture, catgut embedding, and emerging technologies (e.g., laser acupoint irradiation), have been applied clinically with modest success. For instance, Yu Xueping et al. employed Dredging and Regulating Du-Ren Meridians Acupuncture to treat synkinesis[14], while Yang Tianying et al. used Juci technique combined with heat-sensitive moxibustion[15]; both reported therapeutic benefits. However, these approaches lack quantifiable principles for acupoint selection, exhibiting strong subjectivity, arbitrariness, and poor reproducibility. Moreover, prolonged intensive stimulation of affected facial areas fails to specifically address synkinesis, presenting limitations and risks. As facial paralysis with "paradoxical deviation" involves structural alterations and localized tension changes in muscles, high-intensity acupuncture or electroacupuncture on the affected side may cause tissue reinjury, provoking nerve irritation and facial spasms. Alternatively, it could damage regenerating nerve fibers during repair, leading to aberrant regeneration syndromes (e.g., crocodile tears) or adhesion formation during tissue healing, resulting in synkinesis and even facial muscle atrophy/hypertrophy[16, 17]. Furthermore, evaluation systems lack objectivity, relying primarily on subjective scales like House-Brackmann grading and Facial Disability Index (FDI) questionnaires rather than objective quantitative indicators reflecting local muscle contracture severity and biomechanical property changes (e.g., stiffness) [18]. This impedes therapy optimization and mechanistic exploration, particularly limiting the potential of techniques like Canggui Tanxue acupuncture, which requires precise stimulation of specific deep muscle layers[19].

High-frequency ultrasound shear wave elastography (SWE) technology offers a robust solution for precise localization[20, 21]. SWE noninvasively, quantitatively, and reproducibly measures tissue Young’s modulus values, directly reflecting biomechanical properties (e.g., tissue stiffness) and accurately identifying regions of abnormal tissue hardness[6, 22-24]. Additionally, ultrasound imaging clearly visualizes superficial facial muscles, nerve pathways, and vascular structures, providing real-time "visual navigation" for acupuncture. Guided by ultrasound, practitioners dynamically monitor needle tip position and insertion depth during acupuncture, ensuring precise arrival at pathological sites while avoiding critical adjacent structures (e.g., facial arteries, parotid ducts), thereby enhancing efficacy and safety. We conducted SWE examinations on 30 healthy subjects and 30 patients with facial synkinesis following facial paralysis (**Fig. 1**). The results showed that Young’s modulus values of major facial muscle groups in healthy individuals ranged between 6–9 kPa. In patients with synkinesis, the unaffected side exhibited clear perimysium and Young’s modulus values below 10 kPa. Conversely, the affected side showed significantly elevated Young’s modulus values (mean >10 kPa), markedly exceeding the normal reference range (6–9 kPa), accompanied by enhanced perimysial echogenicity and structural blurring in the Levator Labii Superioris (**Fig. 1**a), Depressor Anguli Oris (**Fig. 1**b), and Depressor Labii Inferioris (**Fig. 1**c). Notably, the frontalis and Levator Anguli Oris muscles displayed no such abnormalities on either side. These findings not only indicate that abnormally elevated tension in the Levator Labii Superioris, Depressor Anguli Oris, and Depressor Labii Inferioris is a key pathological feature of synkinesis but also precisely delineate high-tension target sites requiring intervention.

**Fig. 1.**
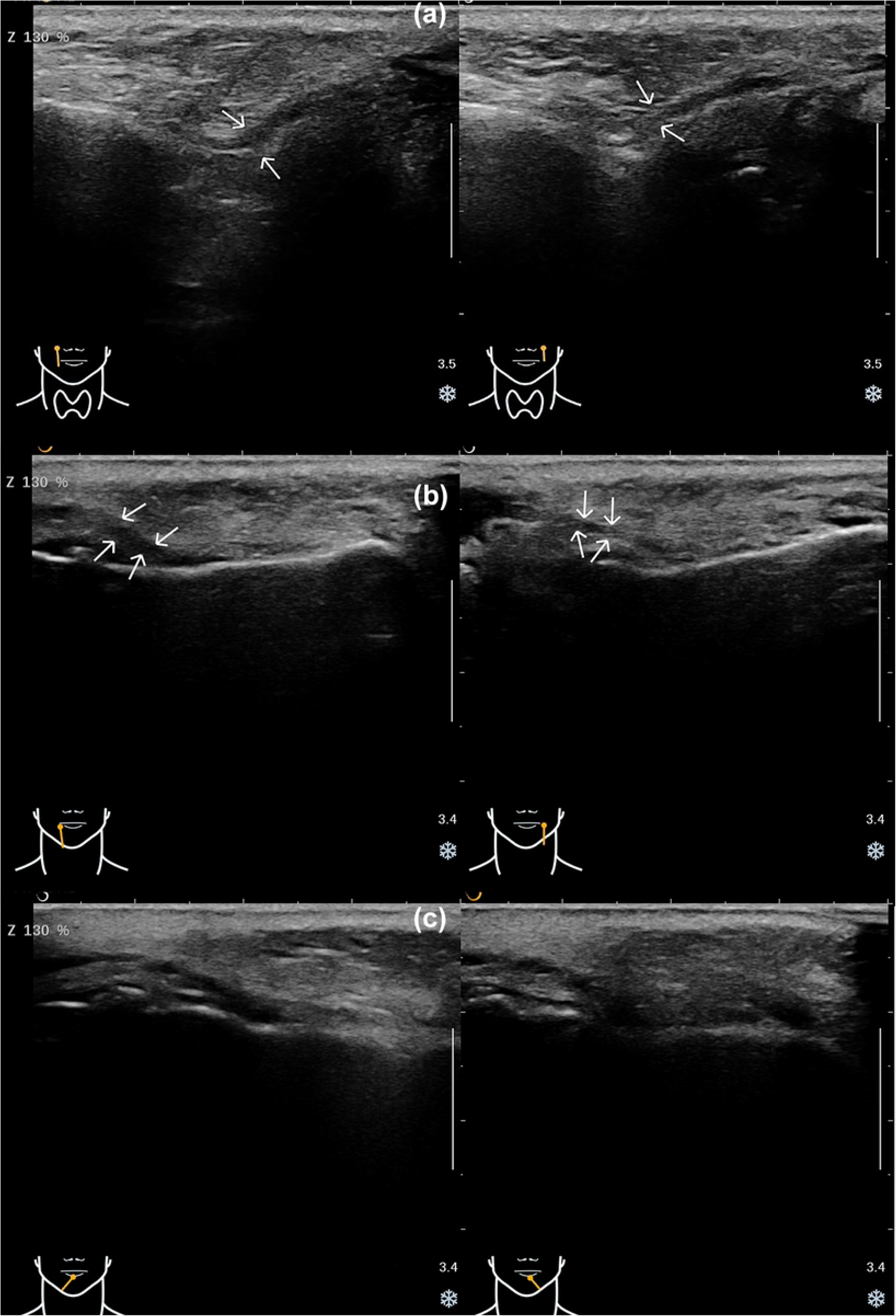
Comparative ultrasound imaging of facial muscles in synkinesis patients. (a) Levator Labii Superioris: Left panel displays healthy side with clear perimysium; right panel shows affected side exhibiting blurred, hyperechoic perimysium. (b) Depressor Anguli Oris: Left panel depicts affected side with blurred, hyperechoic perimysium; right panel presents healthy side with clear perimysium. (c) Depressor Labii Inferioris: Left panel illustrates affected side with blurred, hyperechoic perimysium; right panel demonstrates healthy side with clear perimysium.

Based on these pathological characteristics, SWE’s precise localization capabilities, and the efficacy of Canggui Tanxue acupuncture in releasing deep tissue adhesions, this study designed and implemented a randomized controlled trial (RCT). It objectively evaluated the clinical efficacy and safety of SWE-guided Canggui Tanxue acupuncture for facial synkinesis post-paralysis. Sixty-four eligible patients were enrolled (Control Group: n=33; Ultrasound-guided Group: n=31). The Ultrasound-guided Group received real-time SWE-guided precise acupuncture at sites of abnormally elevated Young’s modulus within the affected Levator Labii Superioris, Depressor Anguli Oris, and Depressor Labii Inferioris, using the Canggui Tanxue technique. The Control Group received conventional acupuncture based on meridian theory combined with the facial "three-line method". Both groups underwent six treatment sessions (twice weekly for three weeks). Efficacy was assessed through multidimensional indicators comprising shear wave elastography (SWE)-quantified changes in Young’s modulus of target muscles pre- to post-treatment (directly reflecting tension improvement), House-Brackmann (H-B) grading for facial nerve functional recovery, and the Facial Disability Index (FDI) evaluating functional activity (FDIP subscore) and psychosocial impact (FDIS subscore), supplemented by clinical effectiveness rates derived from H-B grade improvement. By comparing intergroup improvements in these indicators, this study validates the efficacy of ultrasound-guided Canggui Tanxue acupuncture in reducing target muscle tension, restoring facial nerve function, and alleviating clinical symptoms, while establishing its value as a novel precision strategy for synkinesis.

## 2 MATERIALS AND METHODS

### 2.1 Ethical approval

This study was approved by the Medical Ethics Committee of Chongqing Traditional Chinese Medicine Hospital (Approval No. 2022-ky-36) and registered at the International Traditional Medicine Clinical Trial Registry (ITMCTR2025000783). All procedures adhered to the Declaration of Helsinki. Before participating, all participants received detailed information about the study phases and required written consent for their participation.

### 2.2 Study Population and Design

This study recruited patients with post-paralytic facial synkinesis treated at the Department of Acupuncture, Chongqing Chinese Traditional Medicine Hospital (June 2022-May 2023). Inclusion criteria: Met the 2016 facial neuritis diagnostic criteria of the Chinese Medical Association Neurology Branch with clinical synkinesis; age 16-75 years; time since onset of facial paralysis >3 months; unilateral post-paralytic sequelae with House-Brackmann (H-B) grade ≥II; provided informed consent.

Exclusion criteria: pseudoparadoxical facial deviation; bilateral peripheral/central facial paralysis; paralysis secondary to surgery, trauma, intracranial lesions, or Guillain-Barré syndrome; severe systemic diseases (e.g., diabetes, Cushing’s syndrome), moderate-severe osteoporosis, uncontrolled hypertension, or psychiatric disorders; coagulopathies or bleeding tendencies; pregnancy/lactation.

A 1:1 randomization sequence was generated using SAS 9.4 software. Allocation concealment was implemented using sequentially numbered, opaque, sealed envelopes. As shown in **Fig. 2**, among 67 initially randomized patients, 2 dropped out and 1 was excluded, leaving 64 for final analysis (Ultrasound-guided Group: n=31; Control Group: n=33). Due to the nature of acupuncture, a single-blind design was implemented (outcome assessors blinded to group allocation). Baseline characteristics (sex, affected side, age, disease duration) showed no intergroup differences (P>0.05, Table 1), confirming comparability.

**Fig. 2.**
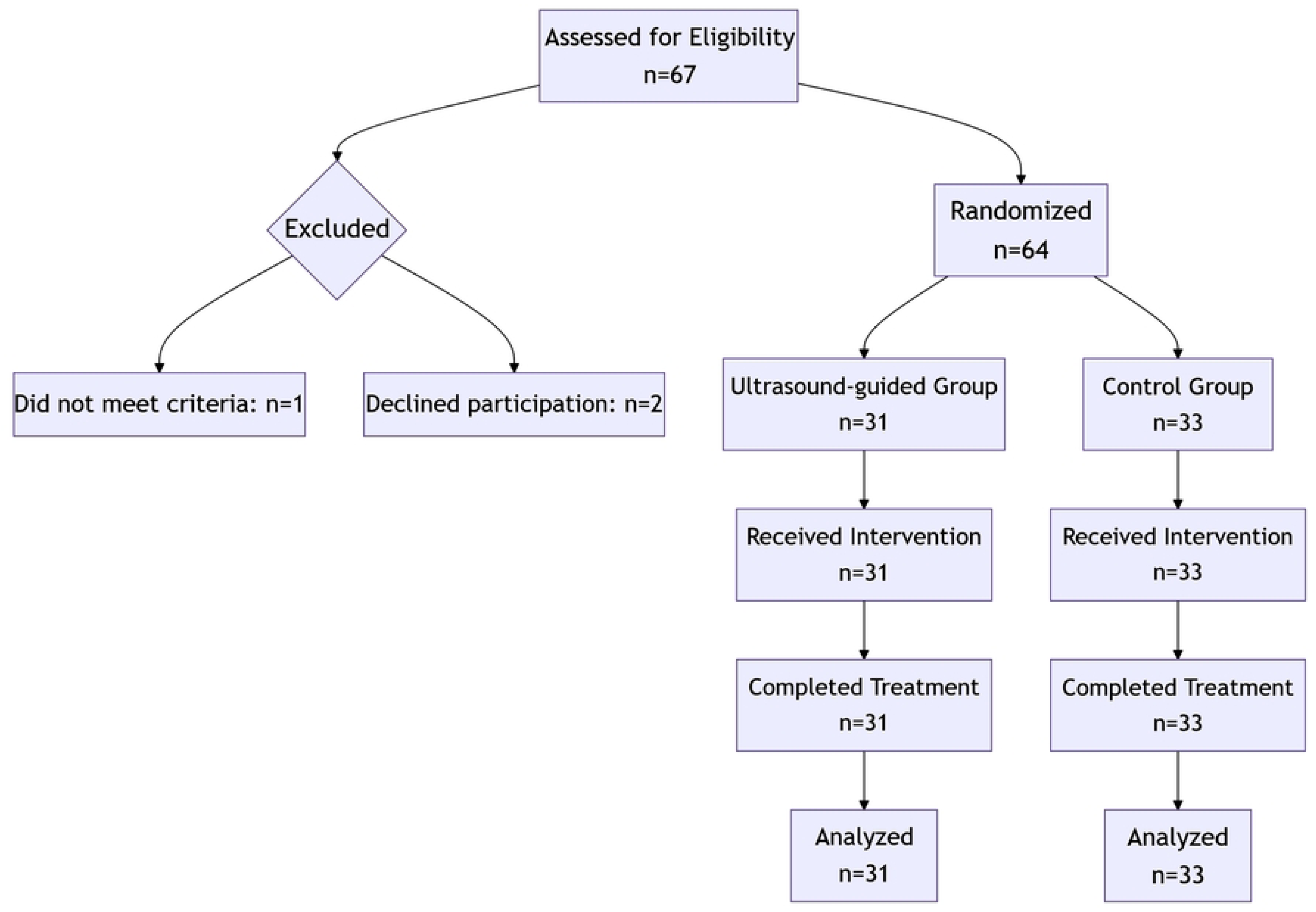
CONSORT flow diagram of participant enrollment, intervention, and analysis. Participant flow: Sixty-seven patients with post-paralytic facial synkinesis were screened for eligibility. Three were excluded (one did not meet inclusion criteria, two declined participation), leaving 64 participants who were randomly allocated to either Ultrasound-guided Group (n=31) or Control Group (n=33). All participants received the allocated intervention and completed the full treatment protocol. Data from all randomized participants were analyzed following the intention-to-treat principle.

**Table 1.**
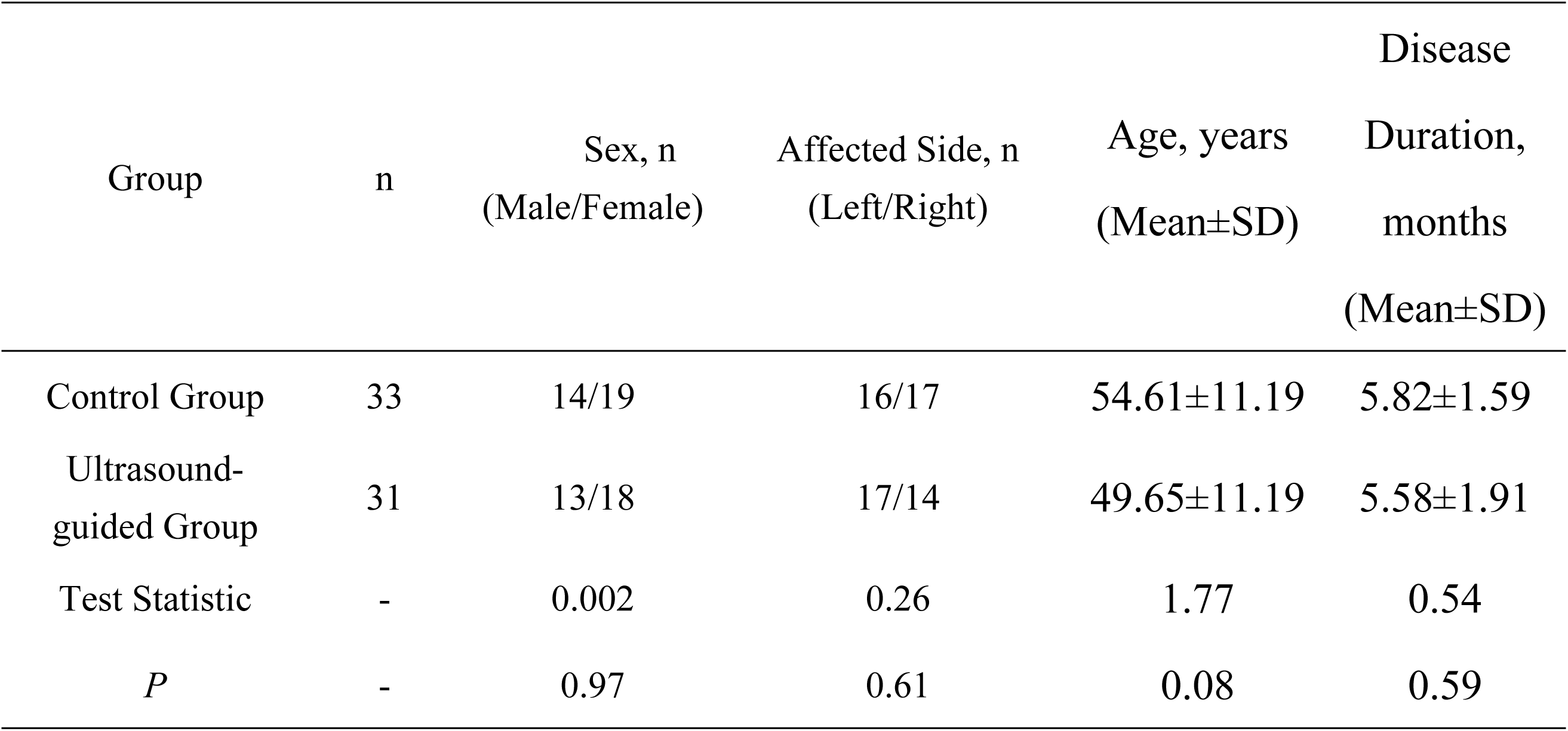
Baseline characteristics of patients with post-paralytic facial synkinesis.

| Group | n | Sex, n<br>(Male/Female) | Affected Side, n<br>(Left/Right) | Age, years<br>(Mean±SD) | Disease |
| --- | --- | --- | --- | --- | --- |
|  |  |  |  |  | Duration,<br>months<br>(Mean±SD) |
| Control Group | 33 | 14/19 | 16/17 | 54.61±11.19 | 5.82±1.59 |
| Ultrasound-guided Group | 31 | 13/18 | 17/14 | 49.65±11.19 | 5.58±1.91 |
| Test Statistic | - | 0.002 | 0.26 | 1.77 | 0.54 |
| *P* | - | 0.97 | 0.61 | 0.08 | 0.59 |

### 2.3 Interventions

Both groups received acupuncture twice weekly for 3 weeks (6 sessions total), administered by the same experienced acupuncturist.

For the Control Group, conventional acupuncture was administered by combining meridian-based acupoints with the facial "three-line method." Primary meridian points included Baihui (GV20), Fengfu (GV16), Taichong (LR3), Hegu (LI4), Shenting (GV24), Taiyang (EX-HN5), Xiaguan (ST7), Yifeng (TE17), Zusanli (ST36), and Neiting (ST44), needled at 0.8–1 cun depth with even reinforcing-reducing manipulation. The three-line facial approach comprised: 1) midline points (Shenting [GV24], Yintang [EX-HN3], Shuigou [GV26], Chengjiang [CV24]); 2) paramedian points approximately 1 cun lateral to midline (Yangbai [GB14], Yuyao [EX-HN4], Chengqi [ST1], Sibai [ST2], Juliao [ST3], Dicang [ST4]); and 3) lateral points (Taiyang [EX-HN5], Xiaguan [ST7], Jiache [ST6]). Supplementary techniques included penetration needling from Yingxiang (LI20) toward inner Yingxiang for nasal dysfunction and Jiachengjiang (EX-HN) for mentalis impairment. Facial needles (0.5– 1.5 cun) were connected to electroacupuncture units delivering continuous wave stimulation for 30 minutes following deqi.

For the Ultrasound-guided Group, Canggui Tanxue acupuncture was performed under real-time high-frequency ultrasound guidance. Prior to each session, pathological sites were localized using a Supersonic Imagine Aixplorer® ultrasound system with S15-4 linear transducer (1-16 MHz). With patients in supine position and facial muscles relaxed, shear wave elastography (SWE) was employed after initial B-mode scanning to identify regions exhibiting abnormally elevated Young’s modulus (>10 kPa) accompanied by enhanced perimysial echogenicity and structural blurring within the affected Levator Labii Superioris, Depressor Anguli Oris, and Depressor Labii Inferioris. A 7-10 mm diameter region of interest (ROI) was placed over target muscles with the elasticity scale preset to 0-460 kPa.

The acupuncturist then performed Canggui Tanxue technique (Video S1): After stabilizing marked target skin with the left thumb, a disposable filiform needle was inserted perpendicularly into the pathological site at 0.5-1.0 cun depth according to facial muscle thickness. Upon achieving deqi, the needle was withdrawn to the superficial layer (without exiting skin) and redirected in superior, inferior, leftward, and rightward directions. Each directional change involved progressive triple-layer insertion (superficial → middle → deep), simulating "a turtle burrowing into soil". Following multi-directional probing, the needle was retained at the deepest layer for 30 minutes. Identical manipulation was repeated at the original site during needle withdrawal. Each pathological target received two complete manipulation sequences per treatment session - once during insertion and once during withdrawal.

### 2.4 Outcome Measures

All outcome assessments were conducted immediately before treatment initiation and after the completion of the 3-week intervention period (6 sessions). The primary efficacy indicator focused on biomechanical changes in affected facial muscles, quantified through high-frequency ultrasound shear wave elastography (SWE) measurements of Young’s modulus (kPa) in the Levator Labii Superioris, Depressor Anguli Oris, and Depressor Labii Inferioris. Improvement magnitude was calculated as the absolute reduction in Young’s modulus (pretreatment value minus posttreatment value), where larger reductions indicated greater alleviation of muscular hypertonicity.

Secondary outcomes encompassed two key clinical domains. Facial nerve functional recovery was evaluated using the House-Brackmann (H-B) grading system, which classifies paralysis severity from Grade I (normal) to Grade VI (complete paralysis), with assessments recorded both pre- and post-intervention. Clinical efficacy was subsequently categorized into four tiers based on H-B improvement: Ineffective (improvement <1 grade), Effective (1-grade improvement), Markedly effective (≥2-grade improvement), and Cured (restoration to Grade I), enabling calculation of the total efficacy rate as [(Effective + Markedly effective + Cured cases) / Total cases] × 100%.

Quality of life impact was assessed via the Facial Disability Index (FDI), comprising two subscales: the Physical Function Score (Facial disability index physical functions, FDIP) and the Social Function Score (Facial disability index social functions, FDIS). The FDIP assessed key symptoms including the difficulty of holding food in one cheek, the difficulty of drinking from a cup, the difficulty of pronouncing specific syllables, the severity of excessive tearing or dryness in one eye, and the difficulty of rinsing the mouth or brushing teeth. These symptoms were rated as None, Mild, Moderate, or Severe, corresponding to scores of 5, 4, 3, or 2 points, respectively. The total FDIP score was calculated as: FDIP = (sum score - 5) × 5, higher scores indicating better function.

The FDIS assessed key symptoms including the duration of feeling calm, the duration of feeling isolated from others, the frequency of losing temper with others, the frequency of waking during sleep, and the frequency of abandoning social activities due to facial paralysis. These symptoms were rated as All of the time, Most of the time, A good bit of the time, Some of the time, A little of the time, or None of the time, corresponding to scores of 6, 5, 4, 3, 2, or 1 point, respectively. The total FDIS score is calculated as: FDIS = (sum score - 5) × 4, lower scores reflecting less psychosocial burden. Assessments using this index were conducted both before treatment initiation and after the completion of treatment.

### 2.5 Statistical Analysis

Data analysis was performed using SPSS 26.0 (IBM Corp.). Continuous variables were first assessed for normality via Shapiro-Wilk testing. Normally distributed data were expressed as mean ± standard deviation (SD) and analyzed using paired t-tests for within-group comparisons and independent t-tests for between-group differences. Non-normally distributed data were presented as median with interquartile range (IQR) and analyzed using Wilcoxon signed-rank tests (intragroup) or Mann-Whitney U tests (intergroup). Categorical data were described as frequencies (percentages) with chi-square tests for intergroup comparisons. Ordinal data (e.g., H-B grades) were analyzed using Wilcoxon signed-rank tests for within-group changes and Mann-Whitney U tests for between-group differences. All statistical tests were two-tailed with significance defined as P < 0.05.

## 3 RESULTS AND DISCUSSION

### 3.1 Biomechanical Improvements Quantified by Shear Wave Elastography

Quantitative high-frequency ultrasound shear wave elastography (SWE) assessments revealed comparable baseline Young’s modulus values across all target muscles between groups (P > 0.05, Table 2). Post-intervention, both therapeutic strategies significantly reduced muscle stiffness in the Depressor Anguli Oris, Depressor Labii Inferioris, and Levator Labii Superioris, with all intragroup comparisons reaching statistical significance (P<0.001; Table 2 and **Fig. 3**).

**Fig. 3.**
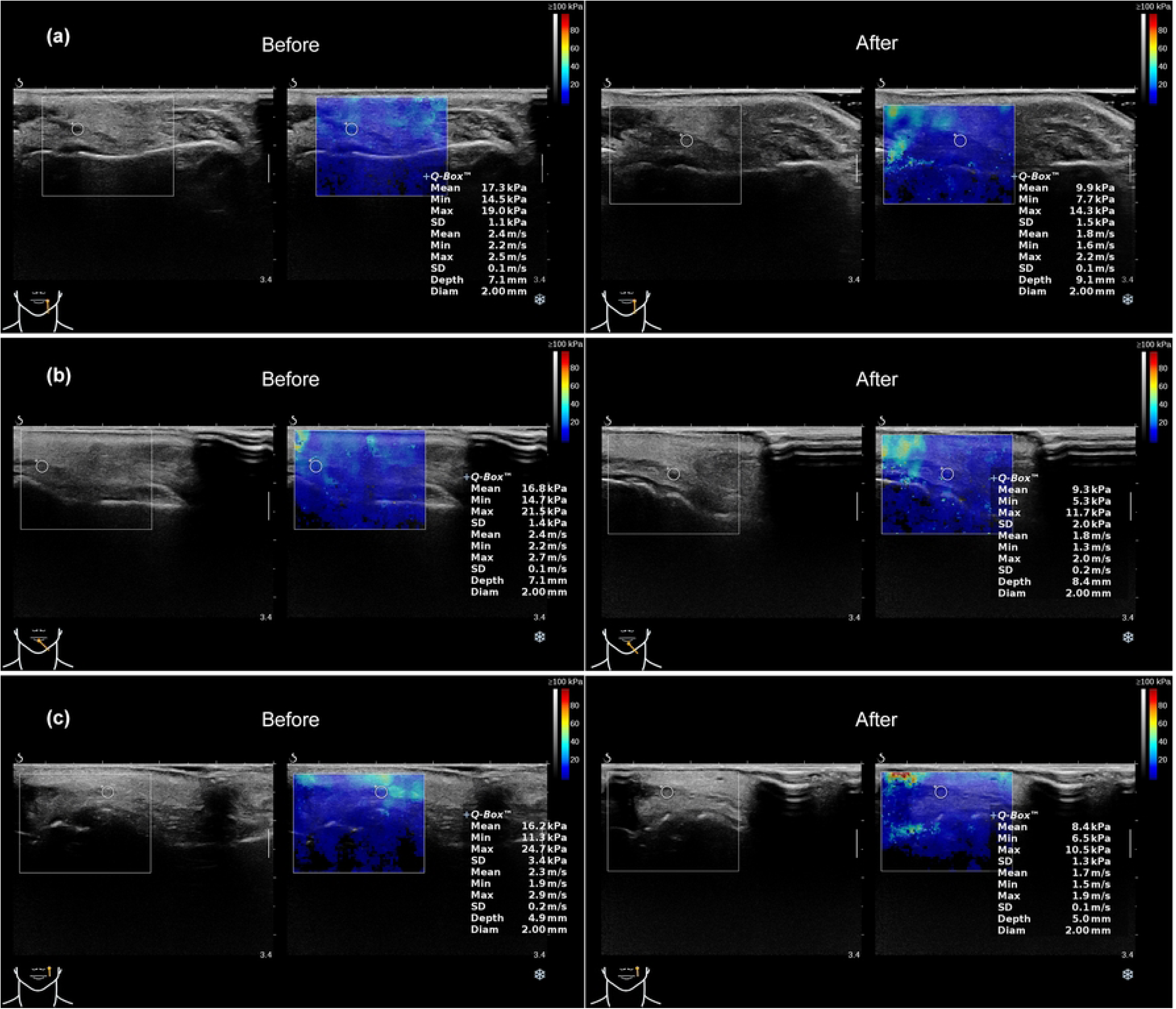
Representative ultrasound and elastographic changes in affected facial muscles of the Ultrasound-guided Group. (a) Depressor Anguli Oris: Left panel (Pretreatment) demonstrates hyperechoic perimysium with blurred architecture and Young’s modulus of 18.2 kPa; right panel (Posttreatment) shows normalized perimysial echogenicity with modulus reduced to 9.2 kPa. (b) Depressor Labii Inferioris: Left panel (Pretreatment) exhibits Young’s modulus of 14.7 kPa; right panel (Posttreatment) reveals reduction to 10.2 kPa. (c) Levator Labii Superioris: Left panel (Pretreatment) displays modulus of 18.6 kPa; right panel (Posttreatment) demonstrates decreased modulus of 13.0 kPa. Dynamic treatment procedure available in Video S1.

**Table 2.**
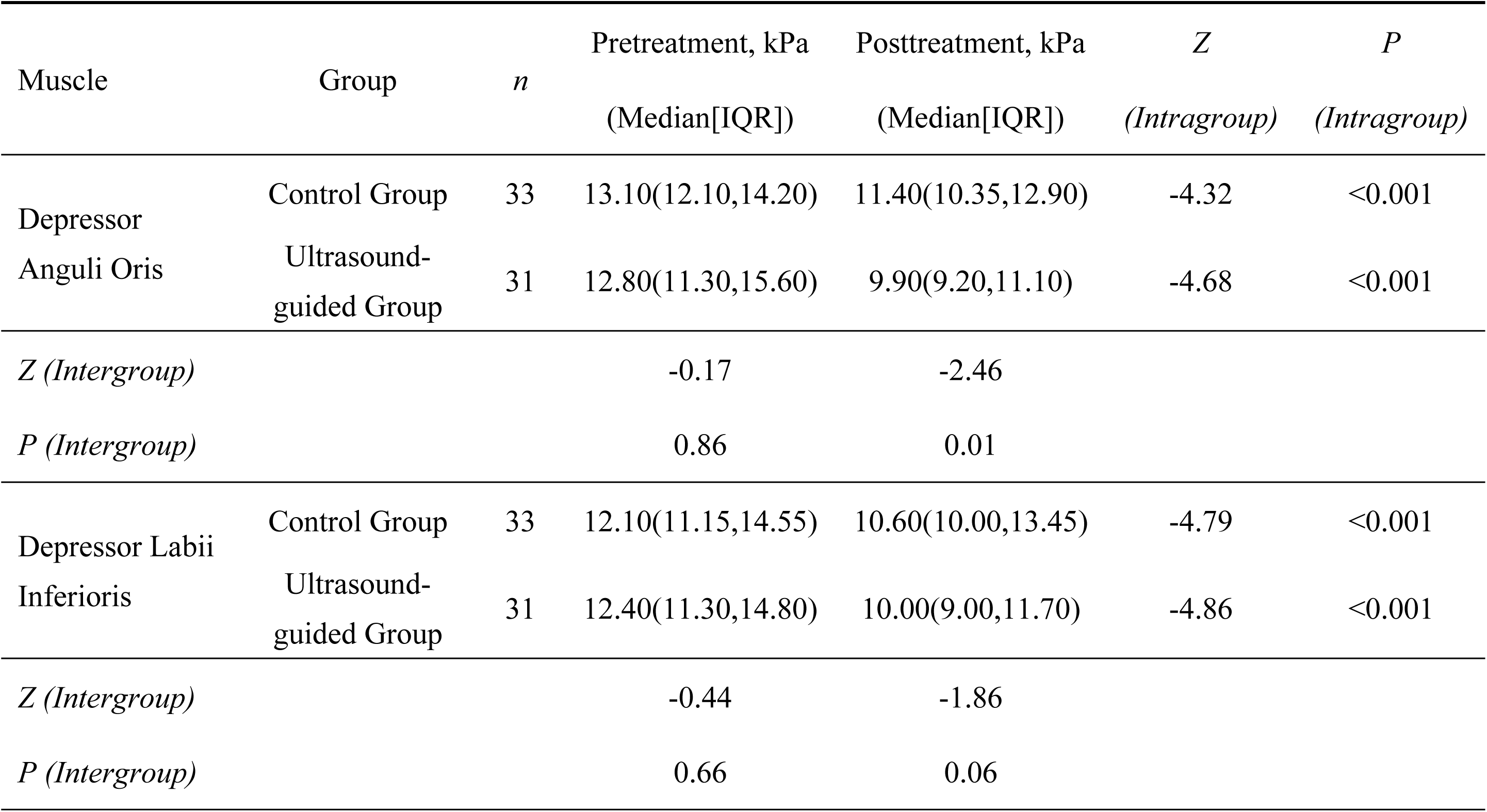
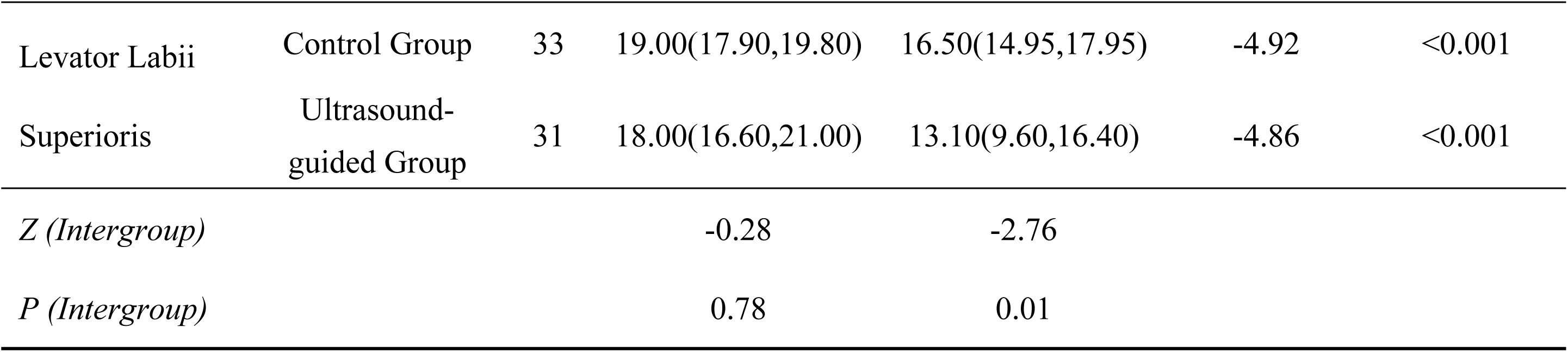
Young’s modulus values (kPa, median [IQR]) in target muscles before and after treatment, and between-group comparisons.

Crucially, ultrasound guidance conferred superior biomechanical efficacy: Depressor Anguli Oris and Levator Labii Superioris exhibited significantly greater modulus reductions in the Ultrasound-guided Group versus Control (median Δ: 3.00 vs. 1.10 kPa, P = 0.001; 5.60 vs. 1.00 kPa, P < 0.001; Table 3). Although Depressor Labii Inferioris showed a clinically relevant reduction trend (median Δ: 2.90 vs. 1.30 kPa), this did not reach statistical significance (P = 0.06), potentially attributable to its deeper anatomical location limiting needle penetration efficacy. The pronounced improvement in Levator Labii Superioris stiffness (**Fig. 3**c, 4c-d) underscores its central role in synkinesis pathology, as hypertonicity in this primary upper lip elevator directly drives oral commissure deviation during voluntary movements. Normalization of tension in Levator Labii Superioris and Depressor Anguli Oris likely rebalances perioral force vectors, facilitating restoration of facial symmetry.

**Table 3.**
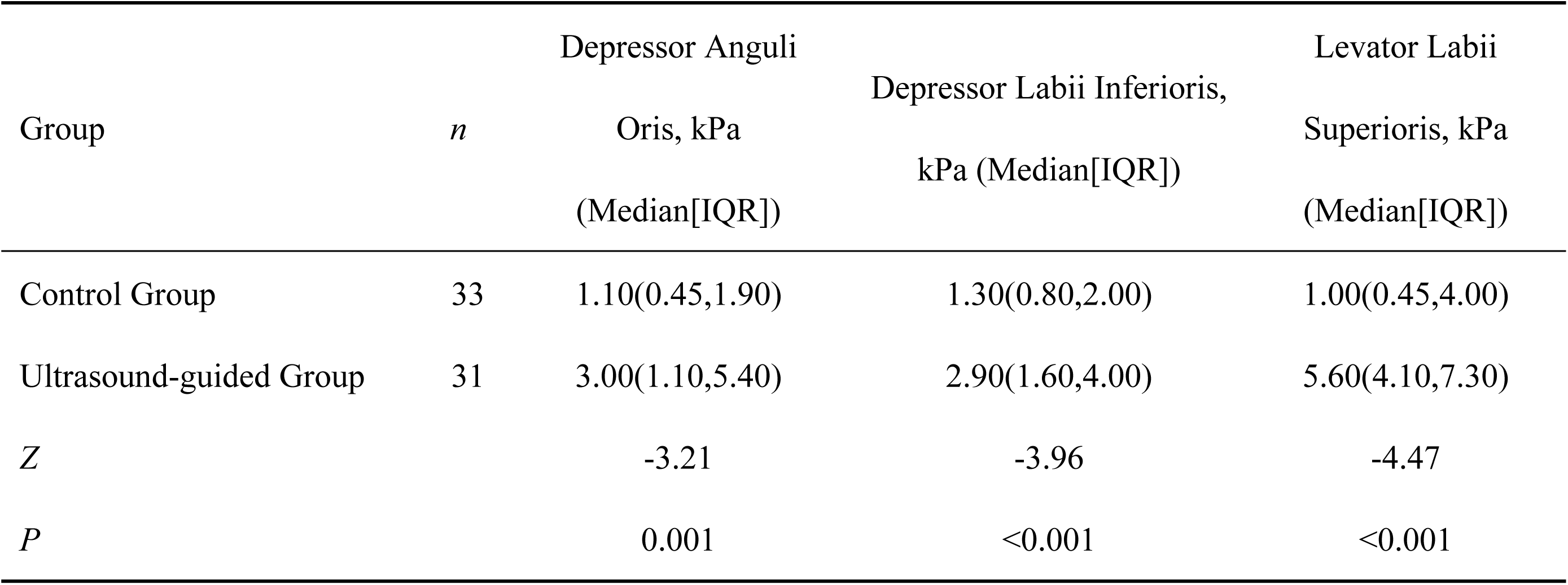
Reduction Magnitude of Young’s Modulus (Δ = Pretreatment - Posttreatment)

**Fig. 4.**
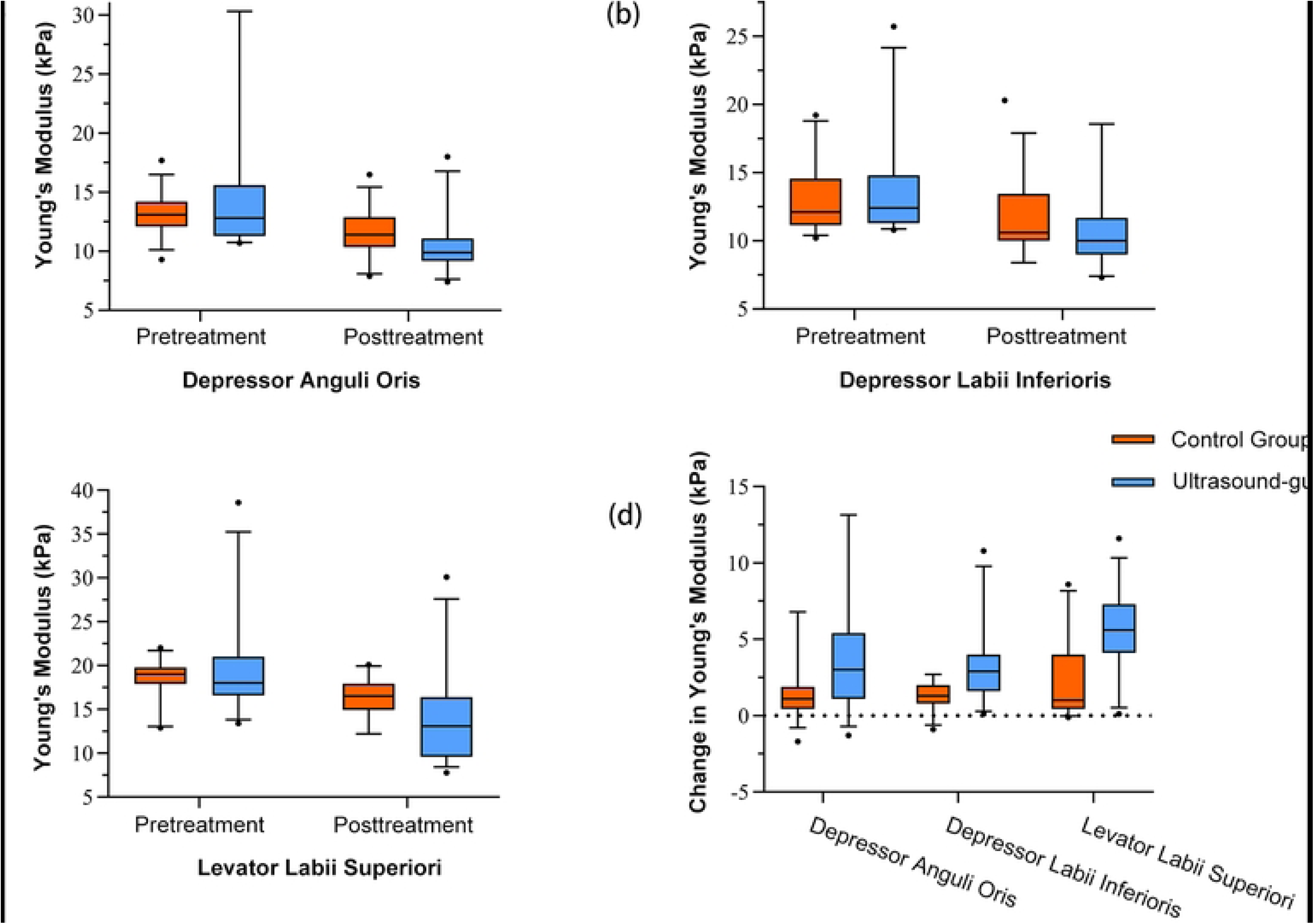
Comparative analysis of Young’s modulus in facial expression muscles. (a) Depressor Anguli Oris modulus values pre- and post-treatment between groups; (b) Depressor Labii Inferioris modulus values pre- and post-treatment between groups; (c) Levator Labii Superioris modulus values pre- and post-treatment between groups; (d) ΔYoung’s modulus (pretreatment-posttreatment difference) for all three muscles between groups. Error bars indicate standard deviation throughout.

### 3.2 Statistical Power Analysis

No a priori sample size calculation was conducted. However, post hoc analysis using the primary outcome data (Young’s modulus reduction in Levator Labii Superioris: 5.6±1.8 kPa vs 1.0±0.9 kPa) showed a Cohen’s d effect size of 3.41. With 31 participants in the ultrasound-guided group and 33 in controls, the study achieved >99.9% power at α=0.05 (two-tailed independent t-test, G*Power 3.1). This exceeds the standard 80% power threshold, indicating the sample size was sufficient for detecting the observed group differences.

### 3.3 Functional Recovery and Clinical Efficacy

Significant improvements in facial nerve function were confirmed in both cohorts via House-Brackmann grading (intragroup P < 0.05), yet ultrasound guidance produced clinically superior outcomes. Post-intervention H-B distribution demonstrated statistically significant divergence (Z = -2.475, P = 0.013), characterized by pronounced stratification shifts (Table 4, **Fig. 5**b): the Ultrasound-guided Group achieved higher representation in mild dysfunction (Grade II: 45.2% [14/31] vs. 21.2% [7/33]) while substantially reducing severe impairment categories, moderately-severe dysfunction (Grade IV) decreased to 6.5% (2/31) versus 15.2% (5/33) in controls, and severe dysfunction (Grade V) was fully eliminated (0% vs. 9.1% [3/33]).

**Fig. 5.**
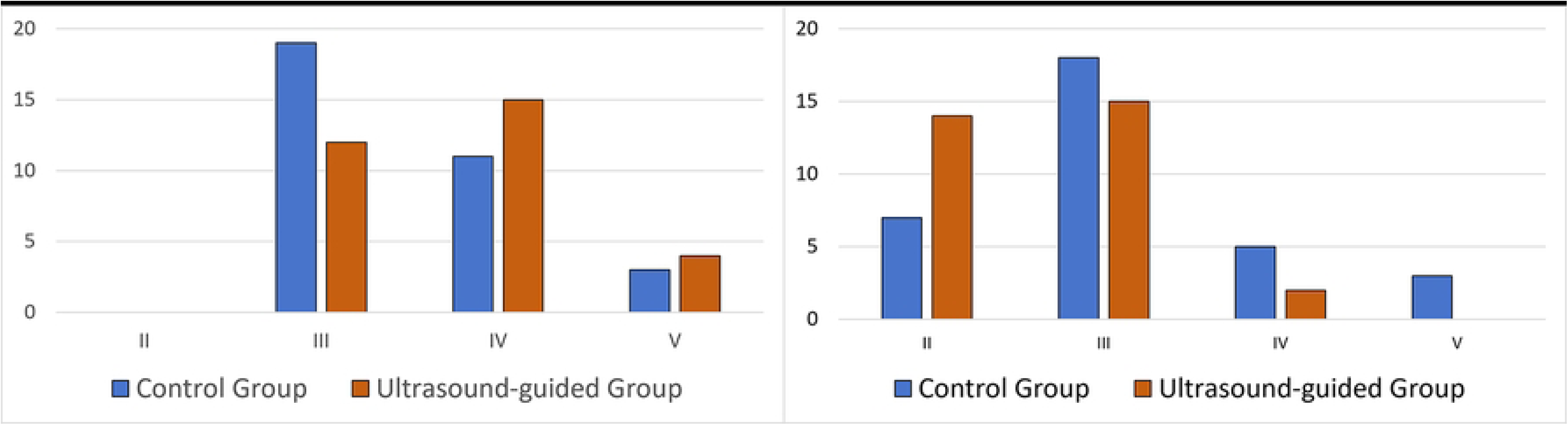
Comparative distribution of House-Brackmann (H-B) grades pre-treatment. (a) Pretreatment distribution; (b) Posttreatment distribution.

**Table 4.** Distribution of House-Brackmann (H-B) grades before and after treatment.

| Time Point | Group | *n* | Grade | Grade | Grade | Grade |
| --- | --- | --- | --- | --- | --- | --- |
|  |  |  | II, n | III, n | IV, n | V, n |
| Pretreatment | Control Group | 33 | 0 | 19 | 11 | 3 |
|  | Ultrasound-guided Group | 31 | 0 | 12 | 15 | 4 |
| Posttreatment | Control Group | 33 | 7 | 18 | 5 | 3 |
|  | Ultrasound-guided Group | 31 | 14 | 15 | 2 | 0 |

This functional restoration translated to a 2.05-fold higher total efficacy rate (87.1% vs. 42.4%, χ² = 13.86, P < 0.001; Table 5). The observed 87.1% total efficacy rate indicates a significant clinical benefit for ultrasound-guided Canggui Tanxue acupuncture in managing post-paralytic synkinesis. This approach may operate through a mechanism potentially involving remodeling of fibrotic tissue architecture, which could yield biomechanical normalization.

**Table 5.**
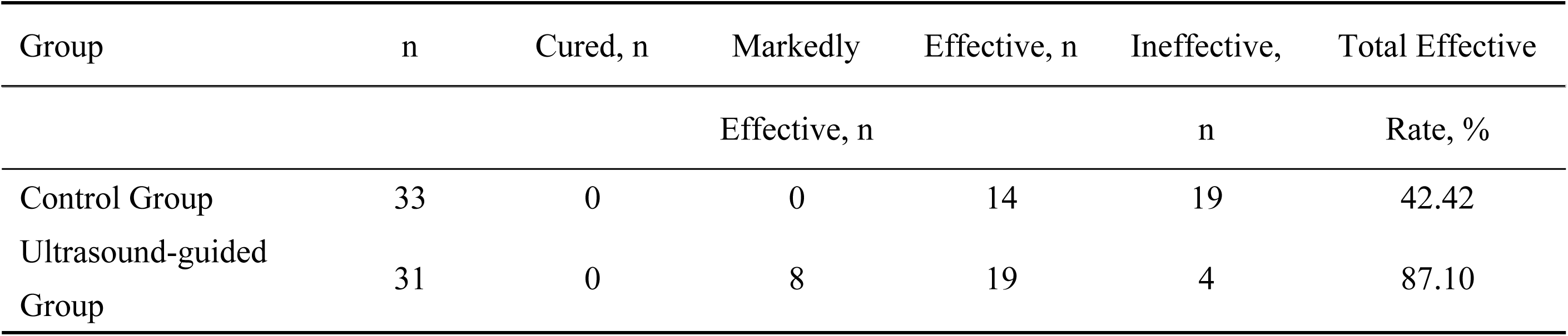
Clinical efficacy rates based on House-Brackmann grade improvement.

### 3.4 Quality-of-Life Outcomes

Physical function (FDIP subscale) improved significantly in both groups (both P < 0.001), yet ultrasound guidance provided superior restoration of daily activities. Final FDIP scores were markedly higher in the Ultrasound-guided Group (74.84 ± 8.42 vs. 68.94 ± 13.68, t = -2.09, P = 0.04; Table 6), reflecting enhanced oral competence during eating and drinking. For psychosocial impact (FDIS subscale), both groups showed reduced scores (both P < 0.001), but intergroup differences were nonsignificant (Z = - 0.10, P = 0.92; Table 7). This discordance suggests that psychological recovery, particularly social anxiety and self-perception, may lag behind functional gains, requiring extended exposure to improved facial control or adjunctive behavioral interventions.

**Table 6.** Facial Disability Index - Physical Function (FDIP) scores before and after treatment.

| Group | n | Pretreatment<br>(Mean±SD) | Posttreatment<br>(Mean±SD) | t Value<br>(Intragroup) | P Value<br>(Intragroup) |
| --- | --- | --- | --- | --- | --- |
| Control Group | 33 | 65.61±13.57 | 68.94±13.68 | -4.30 | <0.001 |
| Ultrasound-guided Group | 31 | 61.13±7.72 | 74.84±8.42 | -10.99 | <0.001 |
| t (Intergroup) | - | 1.64 | -2.09 | - | - |
| P (Intergroup) | - | 0.11 | 0.04 | - | - |

**Table 7.**
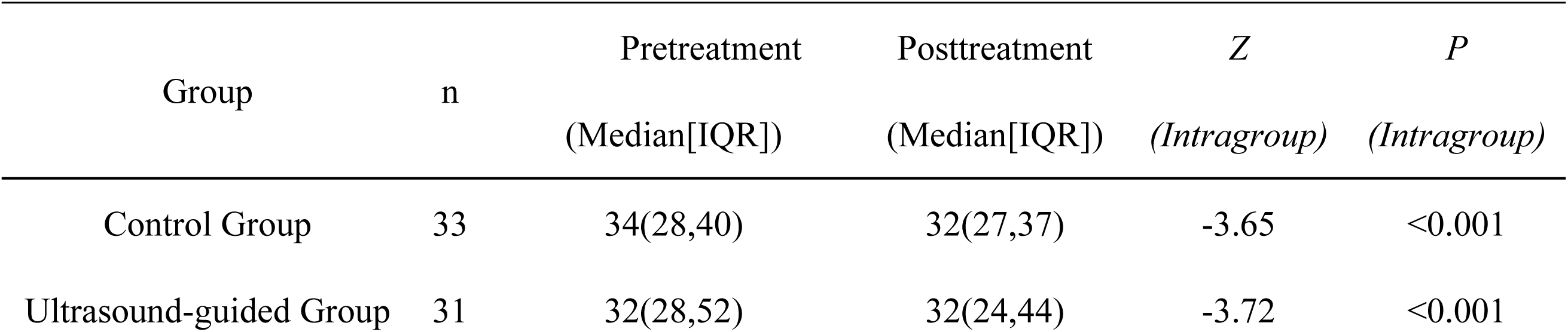
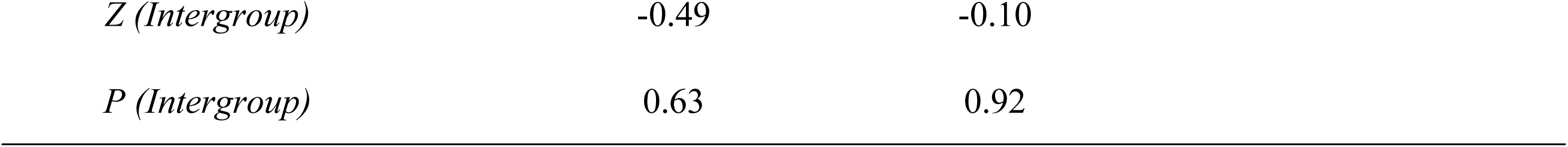
Facial Disability Index - Social Function (FDIS) scores before and after treatment.

### 3.5 Safety Profile

The ultrasound-guided approach demonstrated exceptional procedural safety. Two minor subcutaneous hematomas (6.5% incidence) occurred at needle insertion sites, resolving spontaneously within 48 hours with compression. Critically, no nerve injuries or vascular complications were observed, attributable to real-time visualization of critical structures (e.g., facial arteries, buccal nerve branches) during needle navigation.

### 3.6 Limitations

Several methodological constraints require acknowledgment. First, the exclusive assessment of outcomes immediately following the 3-week intervention precludes evaluation of long-term efficacy, leaving the durability of biomechanical improvements and potential recurrence rates undefined. Second, while shear wave elastography effectively quantified reductions in muscular hypertonicity, the study design did not incorporate neurophysiological investigations to elucidate underlying mechanisms— particularly the relative contributions of peripheral nerve regeneration versus central sensorimotor adaptation to functional recovery. Third, the absence of objective neural metrics, such as electromyographic assessment of axonal integrity or functional MRI mapping of cortical reorganization, limits our ability to establish causal relationships between biomechanical normalization and neuroplastic changes. Future investigations should integrate serial assessments extending ≥ 6 months post-intervention and implement multimodal neuroimaging protocols, including diffusion tensor imaging for facial nerve tract reconstruction and task-activated fMRI for quantifying sensorimotor cortex reorganization patterns.

## 4 CONCLUSION

This randomized controlled trial establishes ultrasound-guided Canggui Tanxue acupuncture as a precise and effective intervention for post-paralytic facial synkinesis. Real-time shear wave elastography navigation enabled targeted modulation of hypertonic musculature (Young’s modulus >10 kPa), yielding significantly greater reduction in pathological stiffness (median Δ: 3.00-5.60 kPa vs 1.00-1.30 kPa; P ≤ 0.001) and superior functional recovery (87.1% vs 42.4% total efficacy rate; P < 0.001) compared to conventional acupuncture at immediate post-treatment assessment. Notably, while physical function (FDIP) significantly improved, psychosocial outcomes (FDIS) showed no intergroup difference (P=0.92), indicating that psychological adaptation may require longer-term intervention beyond biomechanical normalization. The technique demonstrated an exceptional safety profile, with only minor transient hematomas (6.5%) observed and no neurovascular complications. These findings validate the clinical implementation of this biomechanically guided approach. Subsequent research should establish durability beyond the 3-week treatment window and characterize neural remodeling mechanisms through DTI-mediated white matter tractography and fMRI-quantified cortical plasticity metrics.

## Data Availability

All aggregated data necessary to replicate the study's statistical findings are presented within the manuscript (Tables 1-7, Fig. 3-5) and its Supporting Information files. The video demonstration of the ultrasound-guided acupuncture procedure (Video S1) is provided as Supporting Information. Raw ultrasound image datasets generated during this study are prohibitively large. Representative images are included in Fig. 1 and Fig. 3. De-identified raw data for specific cases may be requested from the corresponding author (Jingxin Mao) upon reasonable scientific inquiry, subject to institutional ethics approval.

## Ethical approval

The authors certify that they comply with the Principles of Ethical Publishing. Approval was granted by the Medical Ethics Committee of Chongqing Traditional Chinese Medicine Hospital (ID: 2022-ky-36).

## Author contributions

**Conceptualization:** Jingxin Mao, Yanru Wang

**Methodology:** Yuan Zhou

**Investigation:** Yuan Zhou

**Funding acquisition:** Jingxin Mao, Yanru Wang

**Data curation:** Yuan Zhou

**Project administration:** Jingxin Mao, Yanru Wang

**Resources:** Xiaoming Wu, Xi Zhou

**Formal analysis:** Xiaoming Wu, Xi Zhou, Linjia Wang:

**Supervision:** Linjia Wang

**Writing – original draft:** Yuan Zhou

**Writing – review & editing:** Jingxin Mao, Yanru Wang

## Acknowledgments

This work is supported by Chongqing Science and Health Joint Technology Innovation and Application Development Project (No: 2022MSXM155).

## Supporting information

Video S1. The dynamic treatment procedure guided by ultrasound.

## REFERENCES

1. Husseman J, Mehta RP. Management of Synkinesis. Facial Plast Surg. 2008;24(02):242-9. doi: 10.1055/s-2008-1075840 PMID: 18470836.

2. Terzis JK, Anesti K. Developmental facial paralysis: A review. J Plast Reconstr Aesthet Surg. 2011;64(10):1318–33. doi: 10.1016/j.bjps.2011.04.015.

3. Nellis JC, Ishii M, Byrne PJ, Boahene KDO, Dey JK, Ishii LE. Association Among Facial Paralysis, Depression, and Quality of Life in Facial Plastic Surgery Patients. JAMA Facial Plast Su. 2017;19(3):190–6. doi: 10.1001/jamafacial.2016.1462 PMID: 28122058.

4. Kaiqing L, Tianyang Y, Yuan D, Yuanzheng S, Xue X. Efficacy observation of spirit-regulating acupuncture in treating hemifacial spasm coupled with anxiety and its effect on quality of life Shanghai J Acupunct Moxibust. 2023;42(10):1075–80. doi: 10.13460/j.issn.1005-0957.2023.10.1075.

5. Markey JD, Loyo M. Latest advances in the management of facial synkinesis. Curr Opin Otolaryngol Head Neck Surg. 2017;25(4). doi: 10.1097/MOO.0000000000000376 PMID: 28604403.

6. Vejbrink Kildal V, Rodriguez-Lorenzo A, Pruidze P, Reissig L, Weninger WJ, Tzou C-HJ, et al. Ultrasound-Guided Injections for Treatment of Facial Paralysis Sequelae: A Randomized Study on Body Donors. Plast Reconstr Surg. 2024;153(3). doi: 10.1097/PRS.0000000000010802 PMID: 37285208.

7. Chuang DC-C, Chang TN-J, Lu JC-Y. Postparalysis Facial Synkinesis: Clinical Classification and Surgical Strategies. Plast Reconstr Surg. 2015;3(3). doi: 10.1097/GOX.0000000000000283 PMID: 25878931.

8. Shinn JR, Nwabueze NN, Du L, Patel PN, Motamedi KK, Norton C, et al. Treatment Patterns and Outcomes in Botulinum Therapy for Patients With Facial Synkinesis. JAMA Facial Plast Su. 2019;21(3):244–51. doi: 10.1001/jamafacial.2018.1962 PMID: 30703206

9. Chundury RV, D’Angelo AS, Couch SM, Holds JB. Subjective and Objective Measures in the Treatment of Hemifacial Spasm With OnabotulinumtoxinA. Ophthal Plast Reconstr Surg. 2016;32(2). doi: 10.1097/IOP.0000000000000443 PMID: 25811161.

10. Ton G, Lee L-W, Chen Y-H, Tu C-H, Lee Y-C. Effects of laser acupuncture in a patient with a 12-year history of facial paralysis: A case report. Complement Ther Med. 2019;43:306–10. doi: 10.1016/j.ctim.2019.02.015 PMID: 30935549.

11. Guntinas-Lichius O, Prengel J, Cohen O, Mäkitie AA, Vander Poorten V, Ronen O, et al. Pathogenesis, diagnosis and therapy of facial synkinesis: A systematic review and clinical practice recommendations by the international head and neck scientific group. Front Neurol. 2022;Volume 13 - 2022. doi: 10.3389/fneur.2022.1019554 PMID: 36438936.

12. Mehta RP. Surgical Treatment of Facial Paralysis. ceo. 2009;2(1):1-5. doi: 10.3342/ceo.2009.2.1.1.

13. Sindou MP, Mercier P. Microvascular decompression for hemifacial spasm: Surgical techniques and intraoperative monitoring. Neurochirurgie. 2018;58(1). doi: 10.1016/j.neuchi.2018.04.003 PMID: 29784430.

14. Yu X, Fengling R, Wei Z, Huihui M. Observation on the Therapeutic Effect of Acupuncture for Regulating GV and CV on Prosoplegia Perversion. Henan J Tradit Chin Med. 2021;41(03):437–9. doi: 10.16367/j.issn.1003-5028.2021.03.0101.

15. Yang T, Zhou Z, Tan S. 32 Cases of Facial Paralysis Perversion Treated by Opposing Needling and Thermal Moxibustion. J Clin Acupunct Moxibust. 2013;29(09):42–4.

16. Vargo M, Ding P, Sacco M, Duggal R, Genther DJ, Ciolek PJ, et al. The psychological and psychosocial effects of facial paralysis: A review. J Plast Reconstr Aesthet Surg. 2023;83:423–30. doi: 10.1016/j.bjps.2023.05.027 PMID: 37311285.

17. Lapidus JB, Lu JC-Y, Santosa KB, Yaeger LH, Stoll C, Colditz GA, et al. Too much or too little? A systematic review of postparetic synkinesis treatment. J Plast Reconstr Aesthet Surg. 2020;73(3):443–52. doi: 10.1016/j.bjps.2019.10.006 PMID: 31786138.

18. Yen TL, Driscoll CLW, Lalwani AK. Significance of House-Brackmann Facial Nerve Grading Global Score in the Setting of Differential Facial Nerve Function. Otol Neurotol. 2003;24(1). doi: 10.1097/00129492-200301000-00023 PMID: 12544040.

19. Rongchao Z, Tao W, Binfeng L, Qi L, Kuikui G, Yinan Q, et al. Origin and Exploration of ‘Cang Gui Tan Xue’ Needling Manipulation. J Clin Acupunct Moxibust. 2022;38(08):79–82. doi: 10.19917/j.cnki.1005-0779.022157.

20. Cosgrove D, Piscaglia F, Bamber J, Bojunga J, Correas JM, Gilja OH, et al. EFSUMB Guidelines and Recommendations on the Clinical Use of Ultrasound Elastography.Part 2: Clinical Applications. Ultraschall Med. 2013;34(03):238-53. Epub 2013/04/19. doi: 10.1055/s-0033-1335375.

21. Sande JA, Verjee S, Vinayak S, Amersi F, Ghesani M. Ultrasound shear wave elastography and liver fibrosis: A Prospective Multicenter Study. World journal of hepatology. 2017;9(1):38–47. Epub 2017/01/21. doi: 10.4254/wjh.v9.i1.38 PMID: PMC5220270.

22. Sauer M, Guntinas-Lichius O, Volk GF. Ultrasound echomyography of facial muscles in diagnosis and follow-up of facial palsy in children. Eur J Paediatr Neuro. 2016;20(4):666–70. doi: 10.1016/j.ejpn.2016.03.006 PMID: 27041077.

23. You M-W, Kim KW, Pyo J, Huh J, Kim HJ, Lee SJ, et al. A Meta-analysis for the Diagnostic Performance of Transient Elastography for Clinically Significant Portal Hypertension. Ultrasound Med Biol. 2017;43(1):59–68. doi: 10.1016/j.ultrasmedbio.2016.07.025 PMID: 27751595.

24. Kehrer A, Ruewe M, Platz Batista da Silva N, Lonic D, Heidekrueger PI, Knoedler S, et al. Using High-Resolution Ultrasound to Assess Post-Facial Paralysis Synkinesis—Machine Settings and Technical Aspects for Facial Surgeons. Diagnostics. 2022;12(7):1650. doi: 10.3390/diagnostics12071650 PMID: 35885554.

